# Decentralized Patient-Centric Medical Image Exchange: A Hybrid Blockchain and DICOMweb Architecture

**DOI:** 10.64898/2025.12.29.25343172

**Authors:** Yu Tajima

## Abstract

The fragmentation of medical imaging data across isolated Picture Archiving and Communication Systems (PACS) creates significant barriers to interoperability. This paper presents a functional Proof of Concept (PoC) for a decentralized, patient-centric medical image exchange system. By combining an Ethereum-based smart contract layer for access control and settlement with an off-chain Node.js “Worker” bridge, the system enables automated, peer-to-peer transfer of DICOM studies between disparate Orthanc PACS nodes. We introduce a token-based incentive model to compensate data providers and a cryptographic “Alias” system to preserve patient privacy, complying with recent amendments to Japan’s Next Generation Medical Infrastructure Law. The proposed architecture leverages standard DICOMweb protocols (QIDO-RS, WADO-RS, STOW-RS), ensuring compatibility with existing hospital infrastructure while shifting data sovereignty to the patient. The complete source code is available at https://github.com/cacaobean77-ops/kaken23K14851.

## 1 Introduction

Medical imaging data, primarily stored in the Digital Imaging and Communications in Medicine (DICOM) format, is traditionally siloed within institutional PACS. While internal interoperability has improved, cross-institutional sharing remains administratively burdensome. Patients frequently lack direct control over their records, and the transfer of data for second opinions or referrals often involves physical media or centralized cloud intermediaries that introduce privacy risks and single points of failure.

The primary motivation of this research is to establish a system where patients explicitly approve data access requests via cryptographic signatures, triggering an automated, technical transfer of imaging data. Unlike fully decentralized storage solutions which may face regulatory hurdles regarding data persistence, our hybrid approach maintains data in standard, secure PACS servers (Orthanc) while using the blockchain strictly for access governance and financial settlement.

## 2 Related Work

### 2.1 Blockchain in Healthcare

Blockchain has been extensively explored for Electronic Health Record (EHR) access management. MedRec [1] proposed a decentralized record management system using Ethereum to handle permissions, influencing subsequent visions for a personally controlled health information economy [2]. FHIRChain [3] applies these concepts to HL7 FHIR resources, enabling scalable clinical data sharing. However, these solutions primarily focus on textual or lightweight clinical data and do not fully address the latency and bandwidth requirements of high-resolution medical imaging.

### 2.2 Imaging Interoperability Standards

For imaging exchange, the IHE XDS-I (Cross-Enterprise Document Sharing for Imaging) profile has traditionally defined interoperability [4]. While robust, XDS-I implementations can be complex and centralized. The emergence of DICOMweb (QIDO, WADO, STOW) [5] has provided a lighter-weight, RESTful alternative for web-native imaging applications. SMART on FHIR [6] demonstrates how standards-based apps can run atop EHRs; our work extends this philosophy to the PACS layer.

### 2.3 Hybrid Approaches and IPFS Limitations

Prior work has proposed patient-centric image management frameworks using blockchain [7]. However, many existing proposals rely on IPFS [8] for off-chain storage. While IPFS is promising, it introduces regulatory challenges regarding the “Right to be Forgotten” (GDPR) and data persistence guarantees [9, 10]. Our approach differs by focusing on *DICOMweb-native* transfer orchestration: utilizing existing hospital PACS (Orthanc) for compliant storage, with blockchain serving only as the access signaling layer. This ensures that data remains within legally compliant infrastructure until a valid transfer token is exercised.

## 3 System Architecture

### 3.1 Design Goals

The system was architected with the following core objectives:

- **Patient-Controlled Access**: Data transfer must be cryptographically signed by the patient.
- **Standards Compatibility**: Exclusive use of DICOMweb for search/retrieve/store operations [5, 11].
- **Off-Chain Payload Transfer**: Prevention of blockchain bloat by keeping large binaries off-chain.
- **Economic Incentives**: Implementation of an escrow mechanism to compensate data providers for bandwidth and infrastructure costs.
- **Observability and Governance**: Role-Based Access Control (RBAC) and immutable audit logs.

### 3.2 Architecture

Figure 1 illustrates the system components. The “Worker” acts as a secure bridge between the deterministic blockchain environment and the dynamic hospital intranet.

**Figure 1.**
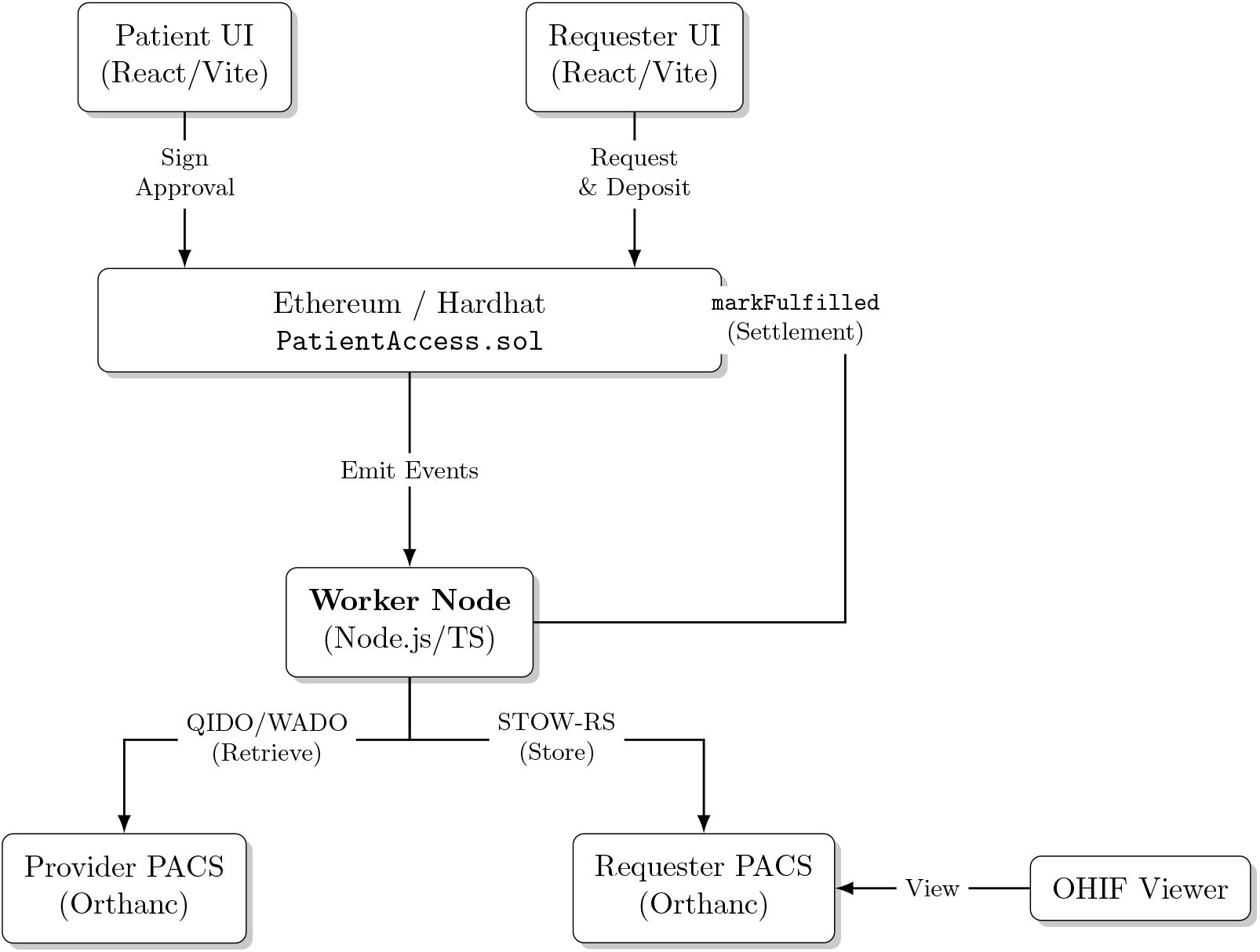
Hybrid architecture diagram. The Blockchain layer governs consent and settlement, while the Worker node orchestrates the off-chain DICOM payload transfer between Orthanc servers. Layout adjusted for clarity.

### 3.3 Smart Contract Interface

The `PatientAccess` contract acts as the state machine and escrow vault. The primary data structure, `AccessRequest`, tracks the lifecycle of a data sharing session. Listing 1 shows the core definitions.

**Listing 1:**
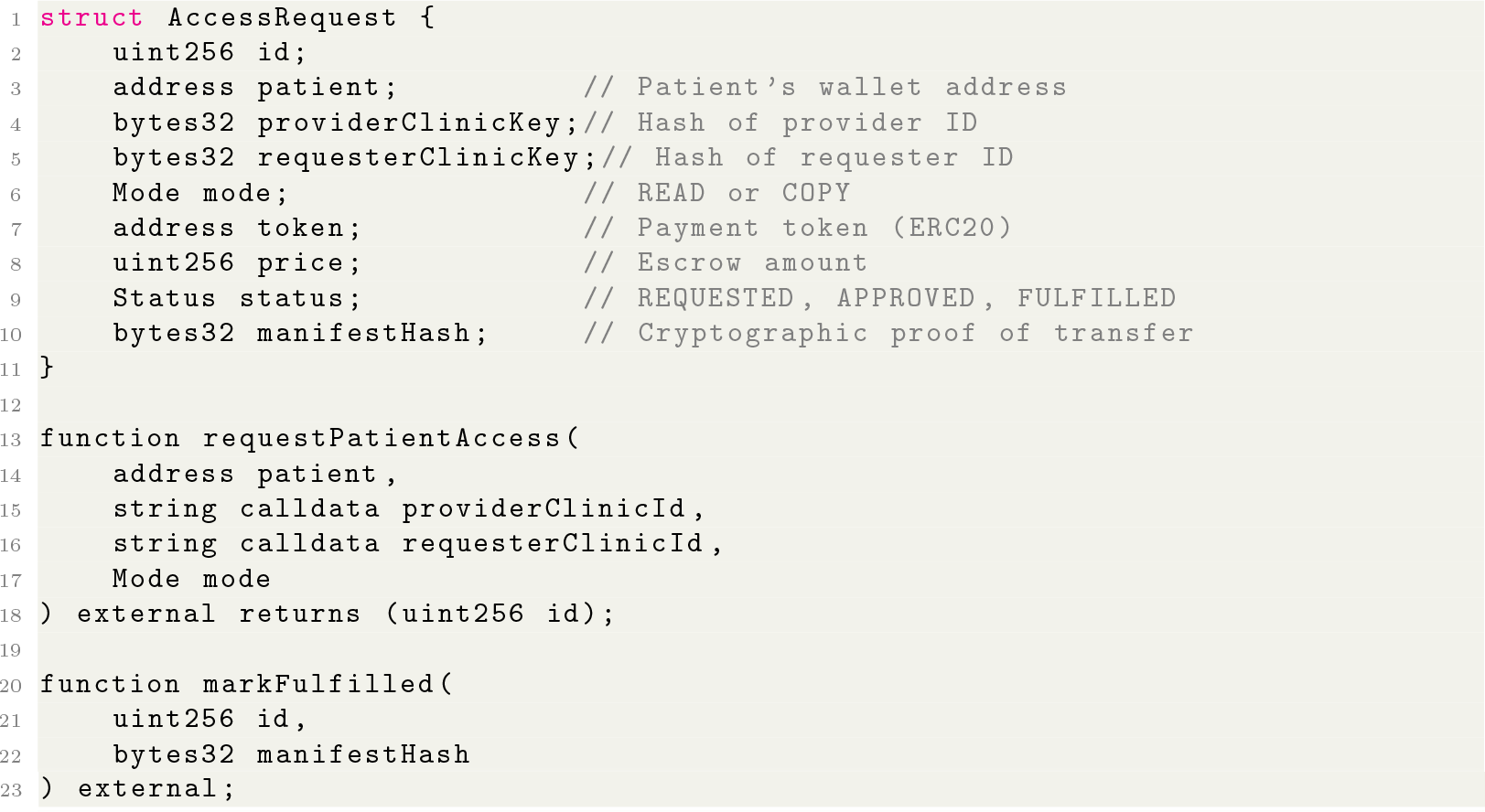
Key data structures and functions in `PatientAccess.sol`

## 4 Methodology and Workflow

The core functionality is the “Copy Workflow,” which facilitates the transfer of a DICOM study from a Provider to a Requester.

### 4.1 Request and Escrow

The Requester selects a patient and specific study types. The Web UI interacts with the `PatientAccess` smart contract to call `requestAccess`. This transaction locks a predefined amount of ERC20 tokens into the smart contract’s escrow balance, creating a financial commitment to the transaction.

### 4.2 Patient Approval

The Patient accesses their portal, reviews the pending request details (Requester identity, purpose of use), and signs a transaction approving it using their wallet (e.g., MetaMask). This emits an `AccessApproved` event on the blockchain, containing the ‘requestId’, hashed ‘patientId’ (Alias), and the ‘providerId’.

### 4.3 Off-Chain Execution (The Worker)

The Worker node, deployed within the Provider’s secure network, listens for the `AccessApproved` event. Upon detection, it performs the following:

1. **Configuration Lookup**: It retrieves the specific Provider’s PACS credentials (URL, Auth) from a local, secure ‘ClinicStore’. This allows for dynamic configuration without restarting the service.
2. **Search (QIDO-RS)**: It queries the Provider’s Orthanc to locate the specific DICOM instances associated with the request.
3. **Transfer (WADO-RS to STOW-RS)**: It retrieves the instances via WADO-RS and immediately pushes them to the Requester’s Orthanc via STOW-RS. This transfer happens entirely off-chain, preserving blockchain bandwidth.

### 4.4 Off-Chain Event Processing

The core logic of the Worker node ensures reliable data transfer by bridging blockchain events with DICOM web services. Algorithm 1 outlines the event handling process implemented in the Node.js service.

#### Algorithm 1

Off-Chain Worker Execution Flow

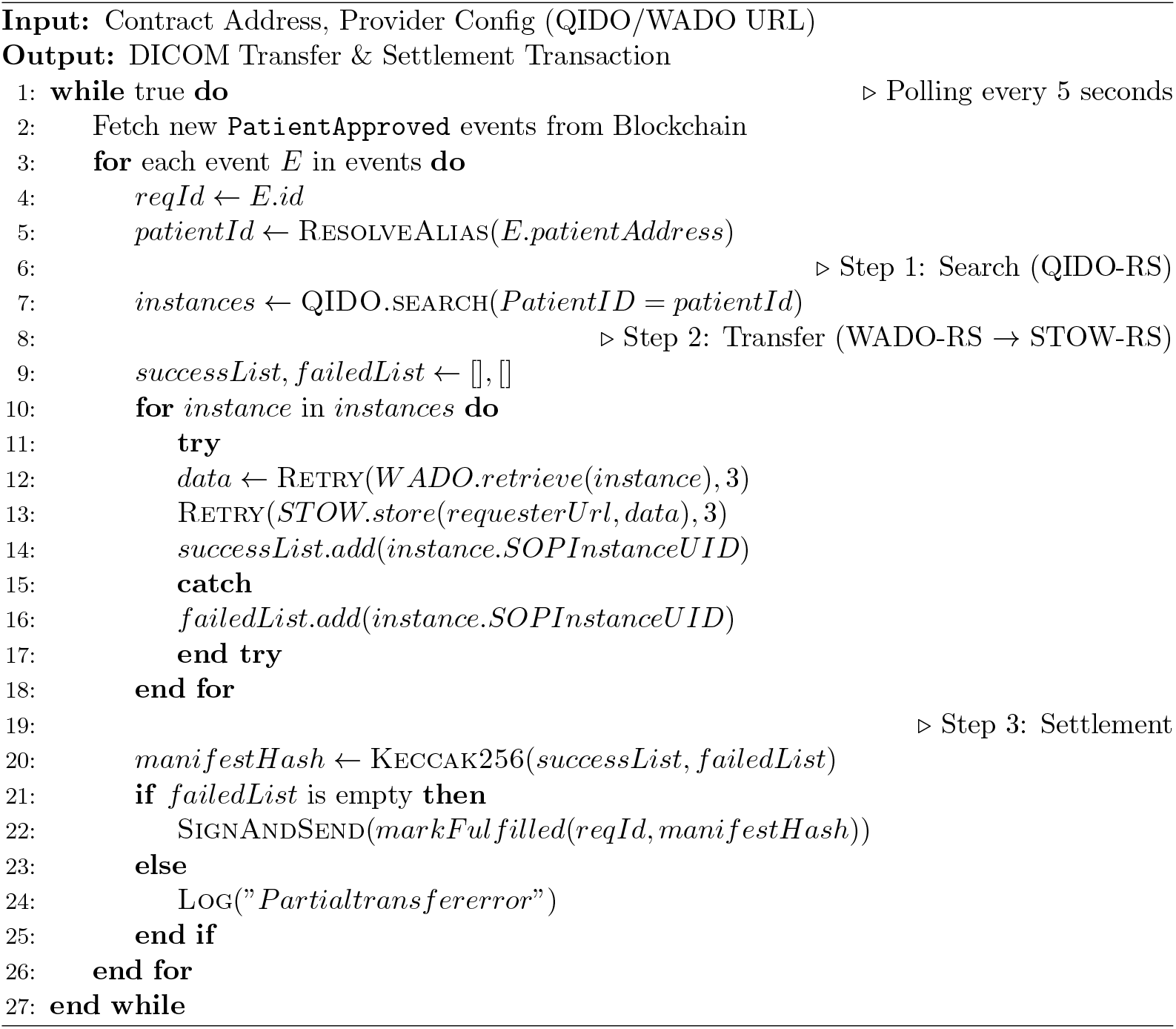

## 5 Evaluation

### 5.1 Gas Consumption and Cost Analysis

To evaluate the economic feasibility of the system, we measured the gas consumption of key smart contract functions. The measurements were conducted using the Hardhat EVM with the optimizer enabled (runs=200). Table 1 summarizes the gas usage and estimated costs on the Ethereum Mainnet versus Layer 2 (L2) networks.

**Table 1:**
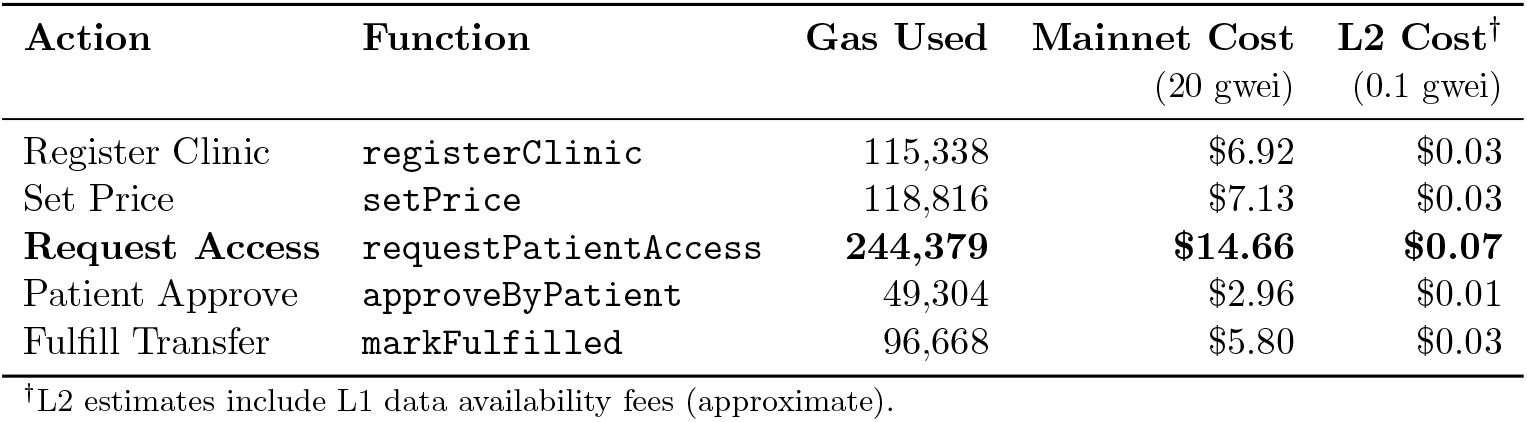
Gas Consumption and Cost Estimation (at $3,000/ETH)

As shown, the `requestPatientAccess` function incurs the highest cost due to the ERC20 token transfer (escrow deposit) and struct storage. On Ethereum Mainnet, a single request would cost approximately $15, which may be prohibitive for routine use. However, on L2 networks (e.g., Optimism, Base), the cost drops to under $0.10, making the system economically viable for frequent medical image exchange.

### 5.1 Performance Latency

The off-chain “Worker” node polls for `PatientApproved` events every 5 seconds. Once an event is detected, the DICOM transfer (Retrieve and Store) is executed via standard HTTP protocols. The latency is dominated by the network bandwidth between the Provider and Requester PACS, rather than blockchain confirmation times.

## 6 Security & Privacy Implementation

### 6.1 Secure Gateway and Access Control

To prevent unauthorized access to the medical images, the Worker node provides a secure reverse proxy gateway (‘/secure/*’). This gateway enforces a strict verification process before forwarding any HTTP requests to the Requester’s PACS:

1. **Token Gating:** The gateway verifies that the incoming request carries a valid JSON Web Token (JWT) with the ‘requester.viewer’ role.
2. **On-Chain Validation:** The gateway queries the smart contract to verify that the ‘requestId’ exists and has a status of at least `PATIENT APPROVED`.
3. **Ownership Check:** It confirms that the requester clinic specified in the smart contract matches the clinic managed by the current Worker instance.

This “Defense in Depth” strategy ensures that even if a viewer URL is leaked, it cannot be accessed without both the off-chain authentication (JWT) and the on-chain authorization state.

### 6.2 Patient Privacy & Alias System

The system complies with the amended Next Generation Medical Infrastructure Law (NGMIL) [12] through an off-chain Alias Manager. Patient wallet addresses are mapped to hospital IDs (PatientID) only within the local Provider environment. On the blockchain, only the hashed alias or wallet address is visible, ensuring that data is mathematically unlinkable to the real-world identity without the Provider’s private mapping key.

## 7 Discussion

### 7.1 Operational Challenges and Viewer Stability

A significant operational finding during the PoC was the instability of web-based DICOM viewers in heterogeneous environments. Specifically, we observed instances where the OHIF Viewer failed to populate the study list correctly (“Study List 0” issue) depending on the browser context. To address this clinical risk, we implemented a multi-tiered viewer strategy:

1. **Primary:** OHIF Viewer for full diagnostic capabilities.
2. **Fallback:** Direct links to the native Orthanc Explorer.
3. **Failsafe:** Display of raw Study Instance UIDs to allow manual retrieval if UI components fail.

This “graceful degradation” approach is critical for maintaining clinical utility even when frontend complexities arise.

### 7.2 Dynamic Configuration Management

In a real-world multi-institutional network, API endpoints and credentials change frequently. Hardcoding these into the smart contract or the Worker image is impractical. Our PoC introduced a ‘ClinicStore’ and a ‘Clinic Config API’, allowing administrators to dynamically update connection details (QIDO/WADO URLs) without redeploying the contract or restarting the Worker service. This feature, combined with Role-Based Access Control (RBAC), allows for the scalable onboarding of new clinics while maintaining security boundaries.

### 7.3 Security and Oracle Integrity

The system mitigates “oracle manipulation” risks by requiring cryptographic signatures from the Provider’s ‘AliasManager’ before any ‘markFulfilled’ transaction can be processed on-chain. Furthermore, the use of standard HTTP-based DICOMweb servers allows for immediate and definitive execution of the “Right to be Forgotten” simply by deleting the file from the Requester’s server, a capability often lacking in IPFS-based solutions.

### 7.4 Future Work: UX and Adoption

Requiring patients to manage cryptographic keys (MetaMask) remains a significant UX hurdle. We identify Account Abstraction (ERC-4337) [13] as a critical next step. This would allow for social recovery mechanisms and gas-less transactions, significantly lowering the barrier to entry for non-technical patients and clinicians.

## 8 Conclusion

We have demonstrated a functional Proof of Concept for a decentralized medical image exchange. By combining the trust and settlement layer of blockchain with the established efficiency of standard medical imaging protocols, the system achieves patient-controlled interoperability without reliance on a central authority. The complete source code is available on GitHub [14] to encourage further research and collaboration.

## Data Availability

All data produced are available online at https://github.com/cacaobean77-ops/kaken23K14851

## Acknowledgments

This work was developed in the context of a Japanese KAKENHI early-career research plan focusing on decentralized architectures for medical imaging exchange [15].

